# Modelling complete dynamics of SARS-CoV-2 pandemics of Germany and its federal states using multiple levels of data

**DOI:** 10.1101/2024.11.11.24317088

**Authors:** Yuri Kheifetz, Holger Kirsten, Andreas Schuppert, Markus Scholz

**Affiliations:** Institute for Medical Informatics, Statistics and Epidemiology, University of Leipzig, Haertelstrasse 16-18, 04107 Leipzig, Germany

**Keywords:** COVID-19, SARS-CoV-2 epidemiologic models, dark figure, parameter heterogeneity, parametrization, extended multi-compartment SIR- type model, Input-Output Non-Linear Dynamical System, Bayesian knowledge synthesis, Machine-Learning, pandemic preparedness

## Abstract

Epidemiological modelling is a key method of pandemic management including that of SARS-CoV-2. New insights into epidemiologic mechanics and new data resources require continuous adaptions of modelling approaches. We here present a revised and considerably extended version of our previous SARS-CoV- 2 model implemented as input-output non-linear dynamical systems (IO-NLDS). We now include integration of age-dependent contact patterns, immune waning, and new data resources such as seropositivity studies, hospital dynamics, variant dynamics, non-pharmaceutical intervention measures and dynamics of the vaccination campaigns.

With this modelling framework, we explain the dynamics of several data resources for the complete pandemics in Germany as well as its 16 federal states. The latter also allows us to investigate the heterogeneity of model parameters in Germany for the first time. To achieve this goal, we extend our estimation approach by constraining variation of parameters among the federal states. This allows reliable estimation of a few thousand parameters using hundreds of thousands of data points.

Our approach can be generalized to other epidemic situations or even other areas of application, thus, supporting general pandemic preparedness.

## Introduction

During the last years, the SARS-CoV-2 pandemic imposed a worldwide high disease burden and there are estimates that it will remain high for an unforeseen period of time. Understanding SARS-CoV-2 induced dynamics at several levels of data is of high importance for a proper risk management including planning of vaccination campaigns, non-pharmaceutical counter-measures, clinical resources and general pandemic preparedness. A plethora of biomathematical models were proposed for that purpose but most of them only describe the pandemics for a limited set of data or a limited time frame [1]. We recently proposed a universal approach to parametrize mechanistic epidemiologic models using multiple, often biased epidemiological or clinical data sets. This approach is based on embedding an epidemiologic model as a hidden layer into an input-output non-linear dynamical system (IO-NLDS, [2]), where the input layer represents factors not known by the model such as changing non-pharmaceutical interventions (NPI), vaccination campaigns or occurrence of new variants. The output layer represents different types of observational data which are linked to the hidden layer via so called data models addressing uncertainty and bias of the observational data in relation to modelled state parameters. Unknown model parameters can be estimated by a Bayesian approach using prior information of parameter ranges derived from different external studies and other available data resources.

With the help of this approach, with our previous model we were able to describe pandemic dynamics of Germany until April 2021 using an age-structured SIR-type model as hidden layer. Several changes in the pandemic situation required an update of the underlying epidemiologic model. These changes comprise for example, (1) different contact behavior of age groups, (2) modelling of final disease states of age groups, (3) new replacement dynamics of variants including the possibility of more than two highly prevalent variants at the same time, and, (4) modelling of age-dependent vaccination efficacy, and, most importantly, immune waning.

Moreover, several new data resources became available or were improved during the pandemic requiring new or updated data models to be linked with the updated epidemiologic model. We integrated, for example, data on the progression of the vaccination and booster campaigns per age group and considered respective differences between German federal states. We collected and included external study data on vaccination efficacy, waning dynamics and booster efficacies as well as seropositivity studies. We also considered the heterogeneity of pandemic dynamics across states allowing estimating state-wise variability of epidemiologic model parameters for the first time.

Finally, we improved our modelling architecture in order to speed up and parallelize data processing and calculations coping with the high-dimensional data and parameter space facilitating our proposed full information approach.

## Materials and methods

### **General** *approach*

We consider an input-output non-linear dynamical systems (IO-NLDS) originally proposed as time-discrete alternatives to pharmacokinetic and –dynamic differential equations models [2,3]. This class of models couples a set of dynamical input parameters such as external influences and factors with a set of output parameters, i.e. observations by a hidden model structure to be learned (named **core mode l**in the following). This coupling represents a hybrid modelling concept, in which deterministic model equations reflecting our mechanistic understanding of the pandemic are combined with empirical relationships of state variables and observational data called **data models** in the following. This represents a major feature of our approach because it allows separating the tasks of epidemiologic model development and addressing data issues such as corrections for incomplete or biased observational data prior to model parametrization.

### Concepts and assumptions of the core model

Our core model is of SECIR type and consists of several sub-models to account for heterogeneity of model parameters and to resemble infection histories. These sub-models are characterized by up to three different attributes namely age group, immune status (**is**), including its waning after vaccination or infection events and virus variant (**vv**).

More precisely, we considered the following categories of these features:

- Five age groups: 1-14 years, 15-34 years, 35-59 years, 60-79 years and ≥80 years
- Ten virus variants: WT (wild type), alpha, delta, omicron BA1, BA2 and BA5, BA.2.75 with BQ.1, XBB, BA.2.86 and KP.3
- Four immune statuses: naïve due to absence of vaccinations or infections (**S**) or shortly after first vaccination (**Vac**0), highly protected by either recent vaccination (**Vac**1) or recovery from a recent infection (**R**1), moderately protected (**Vac**2, **R**2), and weakly protected (**Vac**3, **R**3), see Table 1.

**Table 1.** Qualitative properties of the sub-models reflecting different immune statuses. These properties mirror the immune memory induced by vaccination or last infection event.

| Compartment | Risk of infection | Immune status (see Figure 2) | Risk of severe course of disease (Hospital ward $H$, ICU requirement $C$, death $D$) |
| --- | --- | --- | --- |
| $S, Vac_0$ | highest | Naive | Highest risk for $H$, $C$ and $D$ |
| $Vac_1, R_1$ | small | Protected | No risk for $C$ and $D$, reduced risk for $H$ (40% compared to $S, Vac_0$) |
| $Vac_2, R_2$ | medium | | |
| $Vac_3, R_3$ | high | Waned | Medium risk for $H$, $C$ and $D$ (40% compared to $S, Vac_0$) |

We assume four vaccination statuses to account for the fact that protection against infection is incomplete and wanes much faster than protection against critical disease courses (see Table AA8 in **Appendix A**). Assignment of attributes to compartments is modelled as multidimensional parameter arrays (tensors).

We distinguish three major compartment groups by their assigned attributes: (1) Infectable subjects without previous infection events (**Sc**, **Vac**) carry attribute age, (2) Infectable subjects with previous infection events (**R**) carry attributes age, previous virus variant (one of the ten variants: WT; alpha, delta BA1, BA2, BA5, BA.2.75/ BQ.1, XBB, BA.2.86, KP.3), immune status prior to previous infection (naïve, highly, moderately or weakly protected), and, (3) Infected subjects (**E, I**) with attributes age, virus variant and immune status prior to infection. For the latter, we assume that only **I** is contagious. Compartments **H** and **C** representing patients admitted to hospital ward or ICU are only count compartments. They have the same attributes of their originating compartment **I** and represent a second hidden layer of our model, which is later connected to respective observational data.

In more detail, we make the following assumptions.

1. Infected compartments are those carrying a specific virus variant. This applies for the compartments E, I, H, and C.
2. The latent state **E** comprises infected but non-contagious subjects. This is the transient state between becoming infected and becoming contagious.
3. To model time delays in transitions, we frequently divided compartments into sub-compartments with first order transitions. This approach is extensively used in pharmacological models [4]. It was shown that this approach resembles Gamma-distributed transit times [5].
4. The infected state **I** is the only state assumed to be contagious and is divided into four sequential compartments. There is a single branching for the compartment **I**2, from which patients can proceed either to D (death compartment, representing deaths due to COVID-19) or to **I**3. Finally, the efflux of **I**4 enters **R** 1 representing resolved disease courses.
5. All sub-compartments of **I** contribute to new infections, depending on age, virus variant, and immune status of target subjects.
6. The compartment **I**2 is considered the source of severe disease outcomes comprising treatment at hospital wards **H** or ICU (**C**). These contributions are not modelled by fluxes but as counting respective bed occupancies.
7. The compartment **H** represent disease states requiring hospital ward care. We assume that these patients are not infectious due to isolation. The compartment is divided into three sub-compartments, **H** 1**, H** 2, and **H** 3 to allow comparisons with data of hospital ward bed occupancies**. Rhosp** counts resolved disease courses after hospital ward station care.
8. The compartment **C** represents critical disease states requiring intensive care. Again, we assume that these patients are not infectious due to isolation. In analogy to the compartment of hospital ward treatment, this compartment is also divided into three sub-compartments to mimic disease courses allowing comparing the compartment with data of ICU bed occupancies. **Ricu** counts resolved disease courses after critical state to model cumulative data.

Basic qualitative properties of infectable compartments assigned with different immune statuses are provided in Table 1. Respective transitions are displayed in Figure 1. More details of model compartments and their properties are provided in Table AA1 of **Appendix A** .

**Figure 1:**
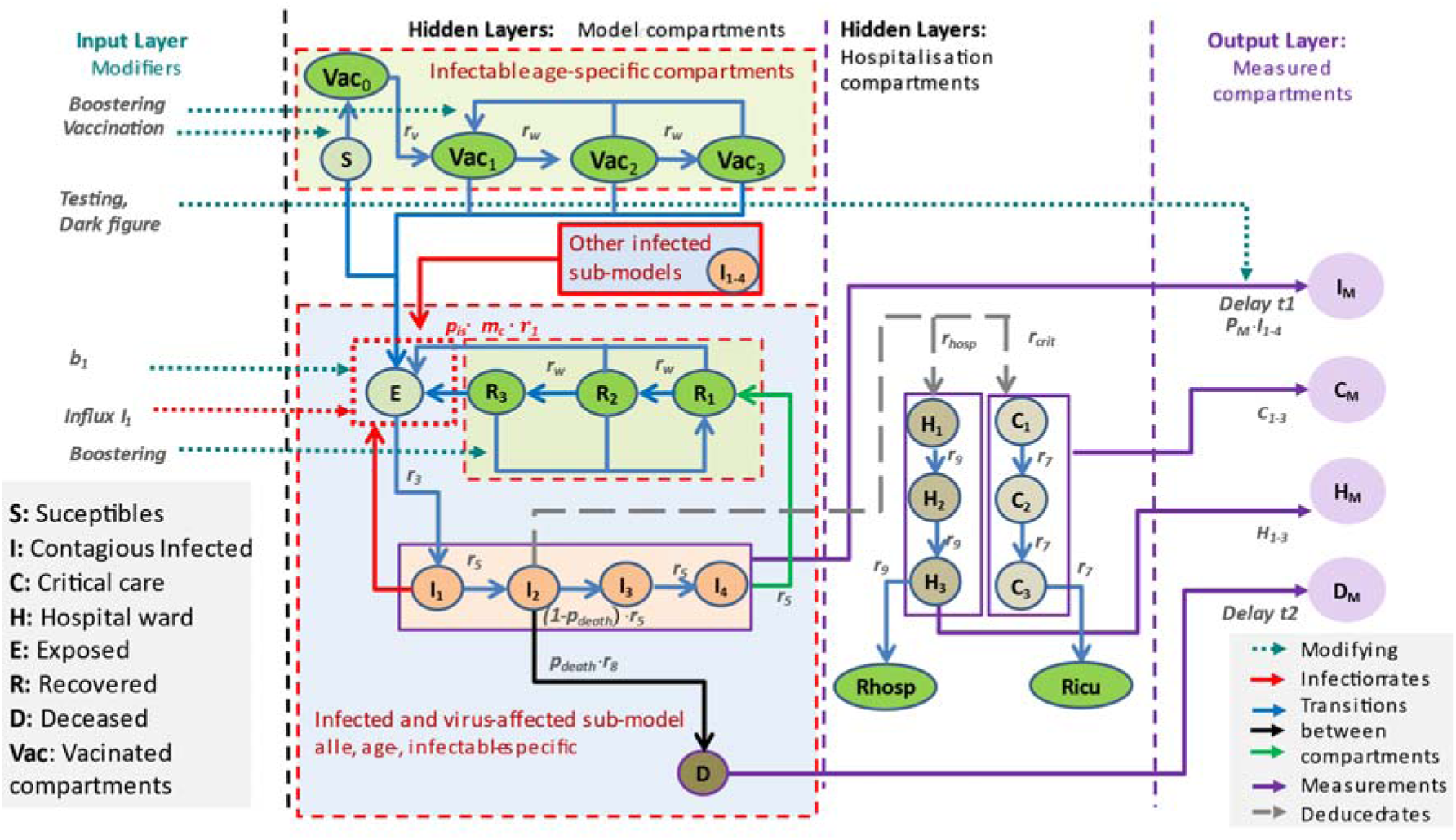
General scheme of our epidemiologic model. Our epidemiologic SECIR model is embedded as a hidden layer into an IO-NLDS. Respective equations are provided in Error! Reference source not found.**B.** Compartments are grouped according to whether they correspond to infectable (green) or infected (blue) subjects. Attributes of sub-models (i.e. *age*, *vv* and and *is*) are not displayed for simplicity. The infectable compartments without previous infection events (S, *Vac*) only depend on age while the other compartments depend on age, virus variant, and if applicable, on the last infection event. The input layer consists of external factors acting on the epidemic such as vaccination campaigns or parameter changes due to changes in testing policy, and non-pharamceutical interventions. The output layer is derived from respective hidden layers via stochastic relationships (*data models*, see later). The output layer is compared with real-world data. The number of hospital (*H*) and ICU (*C*) admissions are described as additional hidden layers counting these events and describing dynamics of bed occupancies.

All model assumptions are translated into a difference equation system (see **Appendix B**). The mathematical structure is that of an Input-Output Non-linear System as depicted in Figure 1. Relationships between immune statuses obtained by vaccination or a previous infection event and possible disease courses are displayed at Figure 2. Model parameters are explained in the **Appendix A** .

**Figure 2.**
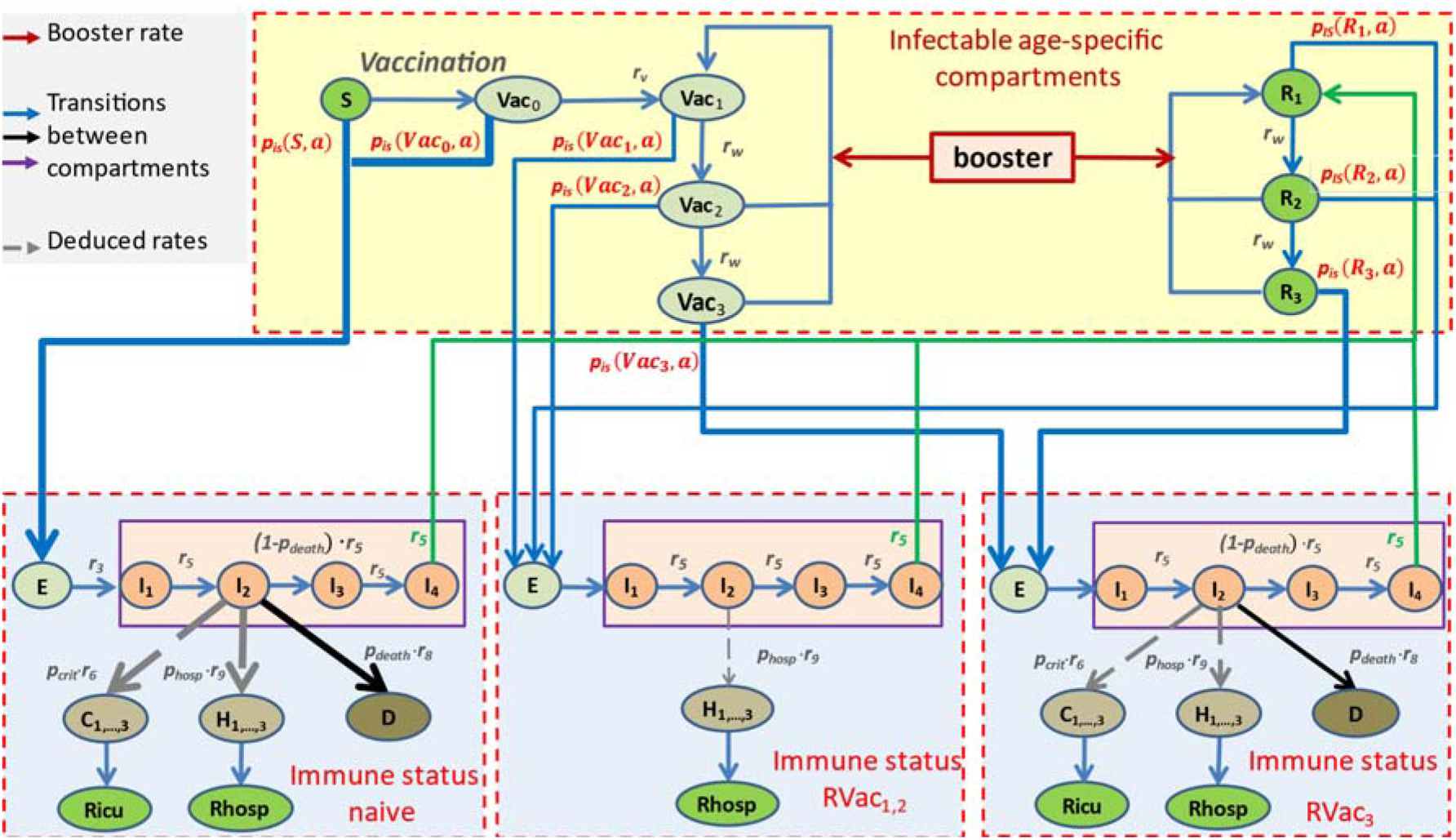
General scheme of the relationships between immune status, infection and severity of disease course. Probability and course of infection depends on the current immune status. Three immune statuses are distinguished: (1) immune-naïve and freshly vaccinated subjects (left), (2) Subjects with recent vaccination or freshly recovered subjects (middle) and (3) subjects with waned immune protection (right). Transition probabilities pIS(Z,a) depend on the current immune status and the infecting variant *a.* Stronger risks are illustrated by broader arrows.

### Input Layer

We here describe the structure of the input layer of our IO-NLDS. This layer is designed to model the impact of external factors acting on the epidemiologic dynamics such as changing infection rates due to non-pharmaceutical interventions, vaccination and booster campaigns and changing testing policies. Effectively, these input functions dynamically affect parameters of the hidden layer containing the epidemiologic model. We describe these different external factors in the following. Respective parameters are described in table 4.

*Dynamical infection rate:* We define a step function b1 as time-dependent input parameter modifying the rate of infections. To identify time points of steps, we used a data-driven approach based on Bayesian Information Criterion (BIC) informed by time points of governmental changes in non-pharmaceutical counter-measures in Germany, changing testing policies as well as events with significant impact on epidemiological dynamics such as holidays or sudden outbreaks [2]. Details can be found in **Error! Reference source not found.** .

*Daily testing and number of undetected cases (estimation of dark figure):* It is well-known that reported numbers of infections are largely underestimated and that this bias is time-dependent during the pandemic. We estimate the dark-figure (DF) based on calibration analyses of seropositivity-data and near-representative systematic testing from the SentiSurv study (See **Err or! R eference sou rce not found.** Respective time-dependent estimates are used for nowcasting true infection numbers to be compared with respective epidemiologic model compartments.

#### Vaccination and booster campaigns

Numbers of applied vaccination and booster doses are available from the German Robert-Koch institute on a daily scale and per age-group and federal state. We distribute respective vaccination rates over eligible model compartments according to their relative size.

#### Output Layer, Data and Parameter Fitting

Unknown parameters of the model are determined by parameter-fitting. For this purpose, compartments of our hidden layer epidemiologic model are coupled with observational data via the output layer of our IO-NLDS using appropriate (stochastic) link functions called **data-models**. We here present these data, respective data-models and objective functions in the following.

We fit our model to age- and federal state-specific time series data of reported numbers of infections **I**M, occupation of hospital stations **N** M, occupation of ICU beds **C** M and deaths **D** M representing the output layer of our IO-NLDS model. Moreover, we fitted data of the variant dynamics.

Data sources of infections, normal ward admissions and deaths were publicly available from the Robert-Koch-Institute (RKI). For our modelling, we used data from March 4^th^, 2020 to September 12^th^, 2024. Number of critical cases, i.e. ICU admissions were retrieved from the German Interdisciplinary Association of Intensive and Emergency Medicine (Deutsche Interdisziplinäre Vereinigung für Intensiv- und Notfallmedizin e.V.—DIVI) for the time window March 25^th^, 2020 to September 12^th^, 2024. Time points in proximity to Christmas and the turn of the year 2020/21 (i.e. December 19^th^, 2020 to January 19^th^, 2021) were heavily biased and therefore discarded during parameter fitting.

Despite this fact, considered data are still largely biased, i.e. cannot be directly linked to state parameters of our epidemiologic model. This was addressed by the following pre-processing steps and data models aiming at removing major biases of

#### Infected cases

**We first smoothed reported numbers of infections with a sliding wind**a**o**p**w**proach **of seven days centered at the time point of interest to remove the strong weekly periodicity of the data. We assume that these numbers correspond to a certain percentage of registered symptomatic patients. To project true infection numbers, we estimate the time-dependent dark figure as explained a**.**b**W**ov**e**e** further account for delays in the reporting of case numbers by introducing a log-normally distributed delay time as explained in Error! Reference source not found..

#### Deaths and hospital ward admissions

Deaths and hospital ward admissions were reported at a daily scale by the RKI since the beginning and end of March 2020 [22], respectively. However, due to data privacy, RKI did not provide exact dates of deaths or hospital ward admissions. Rather than this, reported dates of death and hospitalized patients correspond to the dates of reported infections of these patients. We aimed to remove the resulting reporting delay by assuming log-normally distributed delay times where the expected delay time is derived from the respective transit times of our model. Details can be found in Error! Reference source not found..

Critical cases: Number of critical COVID-19 cases (DIVI reported ICU) was available since the end of March 2020 [21]. We assumed that these data are complete since April 16^th^, 2020 when reporting became mandatory by law in Germany. Earlier data were extrapolated from the number of reporting hospitals using the total number of ICU beds available according to the reported ICU capacity at 2018. These estimates are coupled with the sum of critical sub- compartments **C** i (**i** = 1,2,3) of our model.

#### Frequency of variants

We consider ten virus variants constituting major waves in Germany, namely WT, alpha, delta, omicron BA1, BA2, BA5, BA.2.75/ BQ.1, XBB, BA.2.86 and KP.3. The latter summarizes BA4 and 5 [23]. Entry of new variants is modeled by an instantaneous influx of infected subjects into the compartments E and I [24]. Respective parameters are estimated separately for each federal state. For the purpose of parameter fitting, we also consider data of variant frequencies available for Germany taken from public reports of the RKI [23]. Details can be found in Error! Reference source not found., formula (AE10).

### Parametrization approach

We carefully searched the literature to establish ranges for mechanistic model parameters. We considered two alternatives for parameter estimation: usage of these data as prior information for a Bayesian approach (implemented in our earlier work [2]) or set the parameters from the literature parameters as fixed values (Table A2, justification see Appendix F). Comparison of goodness of fits favoured the second alternative. Parameter estimation is achieved via likelihood optimization. The likelihood is constructed using similar principles as reported previously [25]. In short, the likelihood consists of three major parts, namely a penalty term to ensure that model parameters are within prescribed ranges, as well as penalties for the variability of parameters across federal states as explained in Appendix E. Penalization of variability of parameters across federal states is a sort of pruning to avoid overfitting [26]. We follow a full-information approach intended to use all data collected during the epidemic as explained in Error! Reference source not found.. Consequently, our parametrization approach is intended to describe complete dynamics of the epidemic in Germany and its federal states in the time period covered by the data (Error! Reference source not found. and Error! Reference source not found.).

Likelihood optimization is achieved using a variant of the Hooke-Jeeves algorithm [27].

This is a zero-order algorithm, which does not require expensive calculations of derivatives of the fitness function to be optimized. In brief, the method relies on iterated updates of current fitness values and respective parameter settings by comparisons with fitness values in the neighborhood of the current parameter settings separately for all coordinates. Perturbation sizes at each dimension are adapted in dependence on the result of the previous iteration step, i.e. a perturbation in the s-th dimension becomes larger if a better fitness value was found for this dimension in the previous iteration. Otherwise, it is reduced in the next step, provided that it does not drop below a specified lower limit. An exception are parameters related to the time of entry of new variants for which we used a constant step size of one day in order to stabilize convergence since these parameters are highly sensitive. The algorithm stops if the last four steps did not provide a relative improvement of the fitness function of more than a specified tolerance parameter **δ**tol.

Identifiability of parameters was checked using profile likelihood technique [28]. Details are explained in the supplement. Dynamical parameters were determined by step functions. The number of these steps was determined based on pre-described events or empirically. The Bayesian Information Criterion was applied to penalize the number of steps.

We proved identifiability of key parameters using profile likelihood technique [28]. The profile likelihood of a parameter is obtained when optimizing the likelihood conditional to this parameter: we fix different values of the given parameter and reoptimize all other by maximizing the likelihood. We obtain thus functional dependence of the likelihood function on fixed values of a given parameter. If the fixation of these parameters to the known from the literature values does not lead to improve of the likelihood compared to the case of their estimation, we consider this parameter as redundant. However, this technique is very time consuming. Thus, we applied it to determine identifiability of only few system parameters: the contact matrix terms, relative initial influxes*^age^* and parameters *r*_1_*, r*_3_, *r*_5_, *r*_6,_ *r*_7_ *and r*_9_, for which reliable prior values are known from the literature.

## Implementation

The model and respective parameter estimation procedures are implemented in the statistical software package R from which external publicly available functions are called. The model’s equation solver is implemented as C++ routine and called from R code using the Rcpp package.

## Results

We aim to explain the dynamics of the COVID-19 pandemic regarding infected subjects, hospital ward and ICU occupation, deaths and variant frequency for the entire time period between March 4th, 2020 up to September 12th, 2024 for Germany and its federal states. We first present the results of our parameter fittings. We then show the resulting agreement of model and data for Germany and its federal states and discuss the dynamics of immune statuses. Based on the observation of largely deviating dynamics between federal states, we analyse respective heterogeneity in data and parameters in more detail. Finally, we present a number of validated model predictions.

### Parameter fitting and identifiability

Most of the model parameters were obtained by fitting the predictions of the model to available data of infection dynamics, hospital burden, deaths and variant frequencies. Parameters were set constant based on profile likelihood examination. Overfitting was further controlled by a BIC-based model selection process (see methods).

Likelihood profiling showed that most of the mechanistic parameters of the SECIR model such as the transition rates, and, could be fixed to the values, derived from other studies (Error! Reference source not found.). The likelihood was only sensitive to parameters representing the basic infection rate and representing the hospitalization rate. The results of parameter estimates can be found in Appendix K and Appendix L.

### Comparison of model predictions and observed data for Germany and its federal states

Throughout the pandemic, we observed a good agreement of modelled state variables and their linked output layers representing the data given the underlying data model. Agreement was uniform for the five considered age groups (Figure 3 upper panel) as well as for all 16 federal states (Error! Reference source not found. and Error! Reference source not found.).

**Figure 3:**
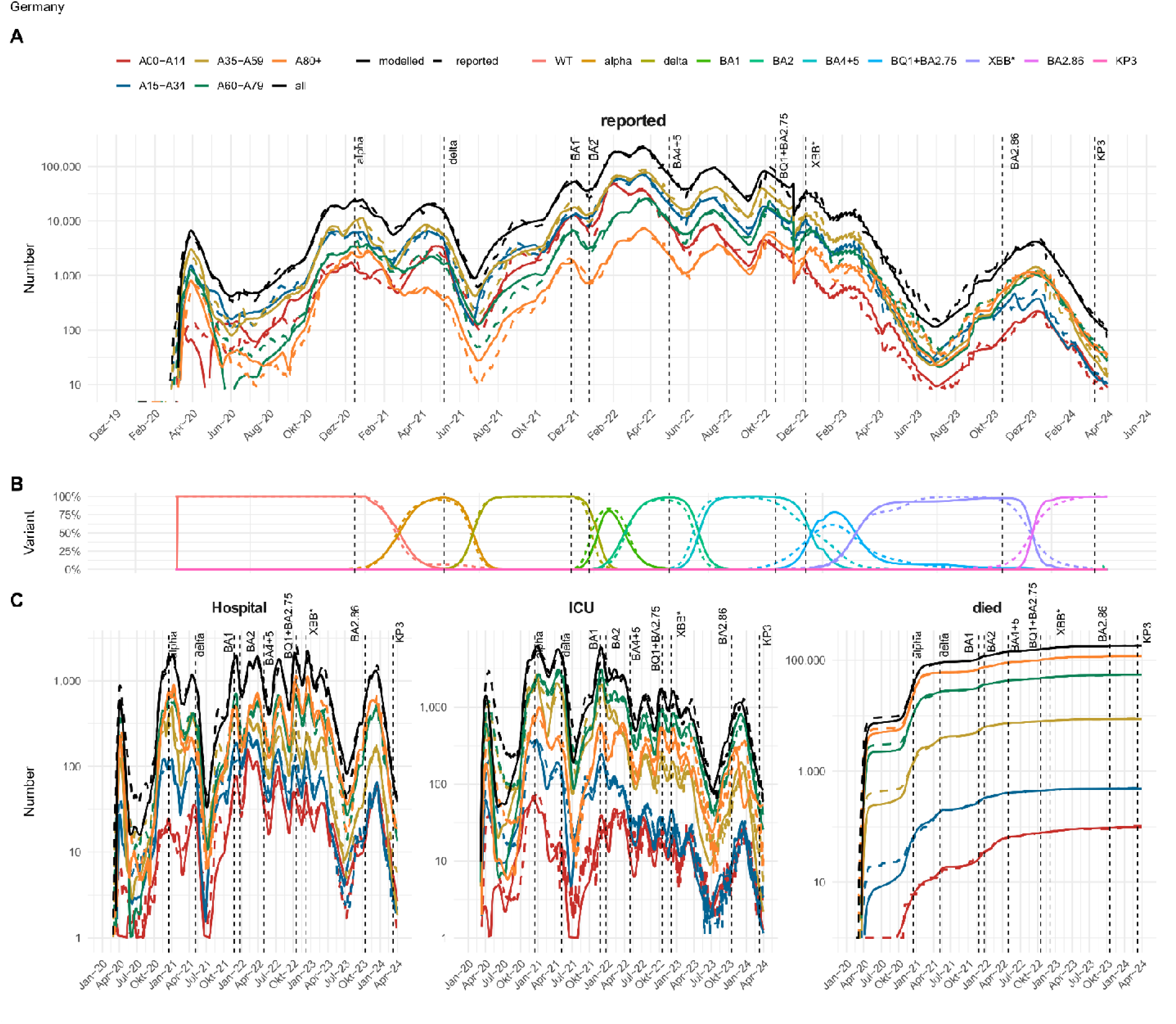
Agreement of model and data for Germany We present the pandemic phase of Germany between March 4th, 2020 and September 12th, 2024 and compare observations (dashed lines) with the predictions of our IO-NLDS model (solid lines). Panel A shows good agreement of model and reported incidence of test positives for all age-groups considered. Panel B shows the dynamical parameter of relative infectivity b1 for the different age groups. Panel C shows the agreement of model and data of variant frequencies. At Panel D we present the model/data comparisons for age-specific dynamics of severe disease states, i.e. hospital ward and ICU occupation and cumulative deaths. Respective figures of the federal states are provided in the supplement.

The model is also in good agreement with the dynamics of virus variant frequencies (Figure 3 lower panel) thereby effectively separating the impact of the variant on the transmission dynamics from other reasons of differing infectivities such as contact behavior or immunity and its waning.

We account for temporarily differing unreported cases specific for age groups and federal counties by estimating a time-variant dark figure based on the reported percentage of positive tests and seroprevalence data. It revealed that the DF is subject to considerable changes in between 50% and 150% unreported infections compared to reported infections for Germany and the modelled time period. DF was even larger for some of the federal states (see Appendix G for details).

Temporal changes of infectivity not explained by virus variants, dynamics of the dark figure and dynamics of immune statuses of subjects is attributed to a stepwise dynamic change of transmission parameter b1 (Figure 3 middle panel). This parameter reflects the strength of contact inhibition at different phases of the pandemic due to NPI or changes in individual contact behaviour, e.g. due to holidays or increased awareness. It is specific for age groups and federal states.

### Dynamics of immune states and their impact on severe disease courses

In our model, (re-)infection probability and susceptibility to severe courses of infection does not only depends on the attacking virus variant but also on the immune states of subjects including their immunization history. Here, we distinguish four different immune states: (1) the immunological naïve state (S/Vac0) with the highest susceptibility to infection and a severe course, (2) high and (3) moderate protection either due to vaccination (Vac1 and Vac2, respectively) or previous infection (R1 and R2, respectively); and (4) low protection due to immune-waning (Vac3, R3). The dynamics of these modelled immune states for Germany is shown at Figure 4, and separately, for the federal states at Error! Reference source not found.. Age-group specific immune states are presented at Error! Reference source not found..

**Figure 4.**
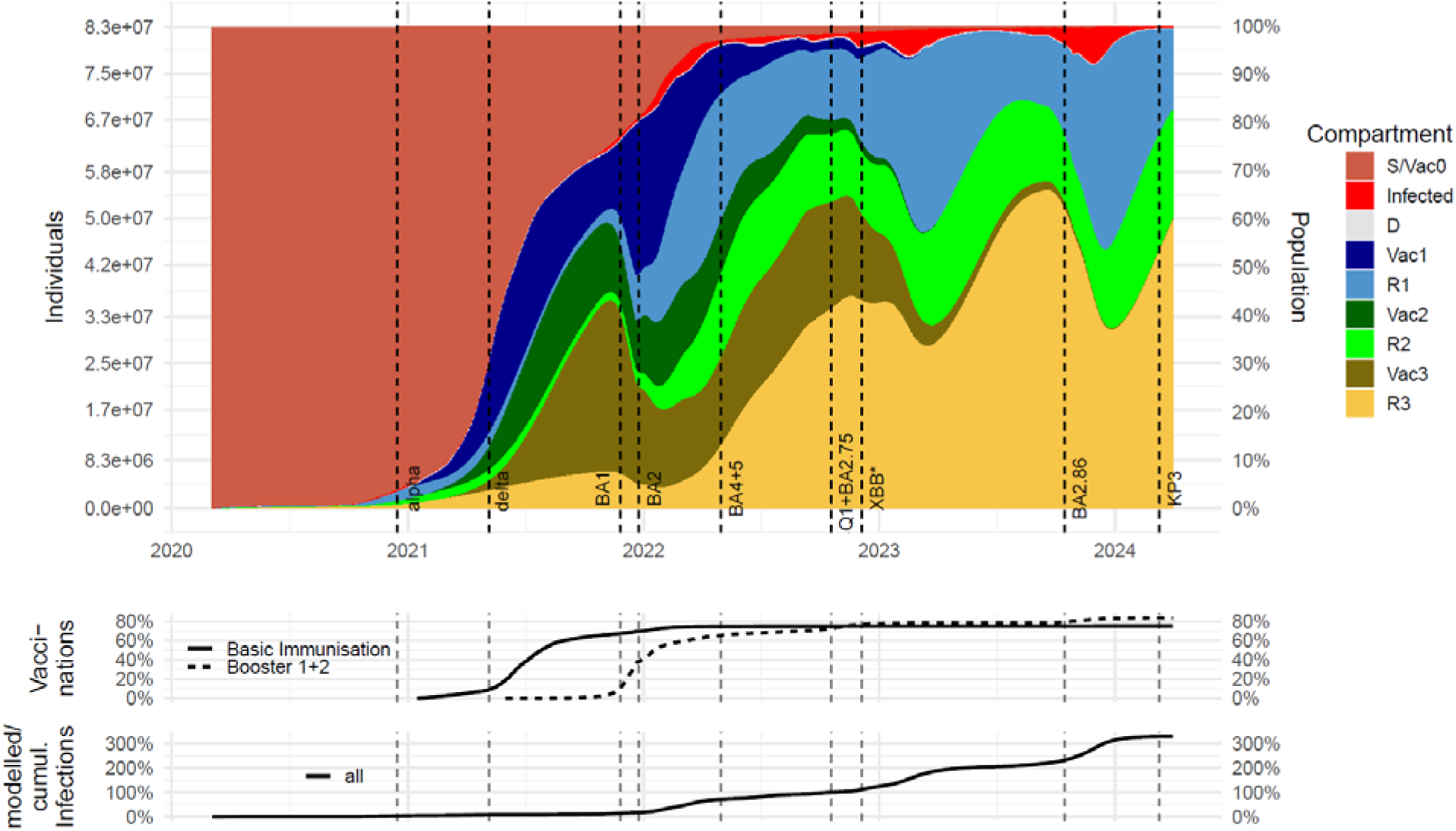
Estimated dynamics of modelled immune states for Germany: We present estimated dynamics of modelled different immune states for Germany (upper panel, S/Vac0 = immune naïve, Vac3/R3 = high risk, Vac2/R2 = moderate risk, Vac1/R1 = low risk). For comparison, we present dynamics of the vaccination campaigns (middle panel) and cumulative number of infections (lower panel).

These dynamics affect the courses of age-specific numbers of hospitalized patients shown in Figure 3D for Germany, and, separated for the federal states, Error! Reference source not found..

### The SARS-CoV-2 pandemic in Germany exhibited strong regional heterogeneity

Epidemic dynamics of federal states differed considerably, which could not be fully explained by difference in vaccination campaigns or age distribution. In this section, we analyse this heterogeneity in more detail by comparing respective data and model results.

We assumed between state heterogeneity for parameters related to severity, i.e. rates affecting the hospitalization (N), ICU (C), and death (D) compartments. We also assumed state-specific parameters related to entries of new virus variants such as timing and initial infection numbers. Moreover, dynamics of the residual infectivity b1 was assumed state-specific. Selection of state-specific parameters was again based on BIC. Of note, it turned out that no differences in viral properties need to be assumed but that the majority of state-specific parameters correspond to population structure and pandemic management, which indeed was heterogeneous across states. During fitting of state-specific parameters, we penalized deviations from the estimates obtained for Germany.

We first analysed the heterogeneity with respect to infection numbers, deaths and test-positivity (see Figure 5A-C). For example, Saxony had a death burden four times higher than that of Schleswig Holstein. Testing policy varied across regions, as evidenced by differences in test positivity rates of up to 75%. Moreover, DF estimates different considerably during the pandemic and between federal states.

**Figure 5:**
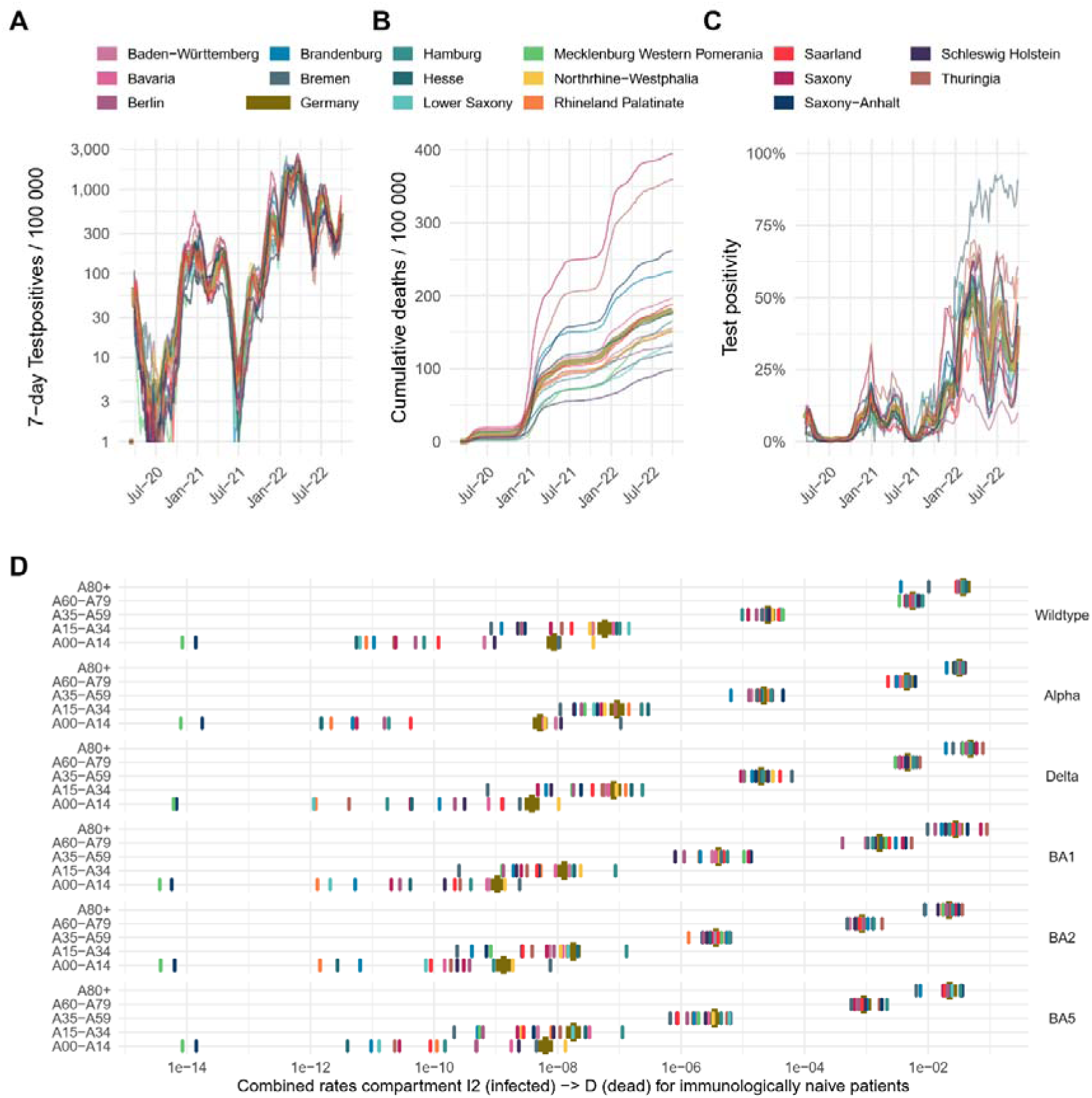
Heterogeneity of the SARS-CoV-2 pandemics between federal countries and region-specific parametrisation of the model. Considerable between-state differences were observed regarding the course or the pandemic examplarily shown by the dynamics of infected subjects (A), reported total number of deaths (B), and probability of test-positivity reflectign testing policy (C).

Estimates of differences of the rates to develop a severe course are shown in Figure XXX and appeared to be plausible: Exemplarily, federal countries Saxony and Thuringia show highest parameter estimates for transition rates to the compartment D (death) in the oldest age group 80+ during the Delta and Omicron-BA1 wave (Figure 5D). Correspondingly, both regions are reported to have suffered from high excess mortality in the elderly. Furthermore, death-rates of Omicron variants are consistently lower than from previous variants (Figure 5D), again in line with the literature. According to our findings, disease dynamics vary across states primarily because of variations in the timing and magnitude of variant introductions, differences in infectivity, which are likely contact-intensity-driven, and differences in severity parameters.

In Figure 6, we compared the between-state heterogeneity of infection dynamics with test positivity and residual infectivity estimates. We also estimated the number and timing of successive introductions of SARS-CoV-2 virus variants in each federal state (Figure 7).

**Figure 6:**
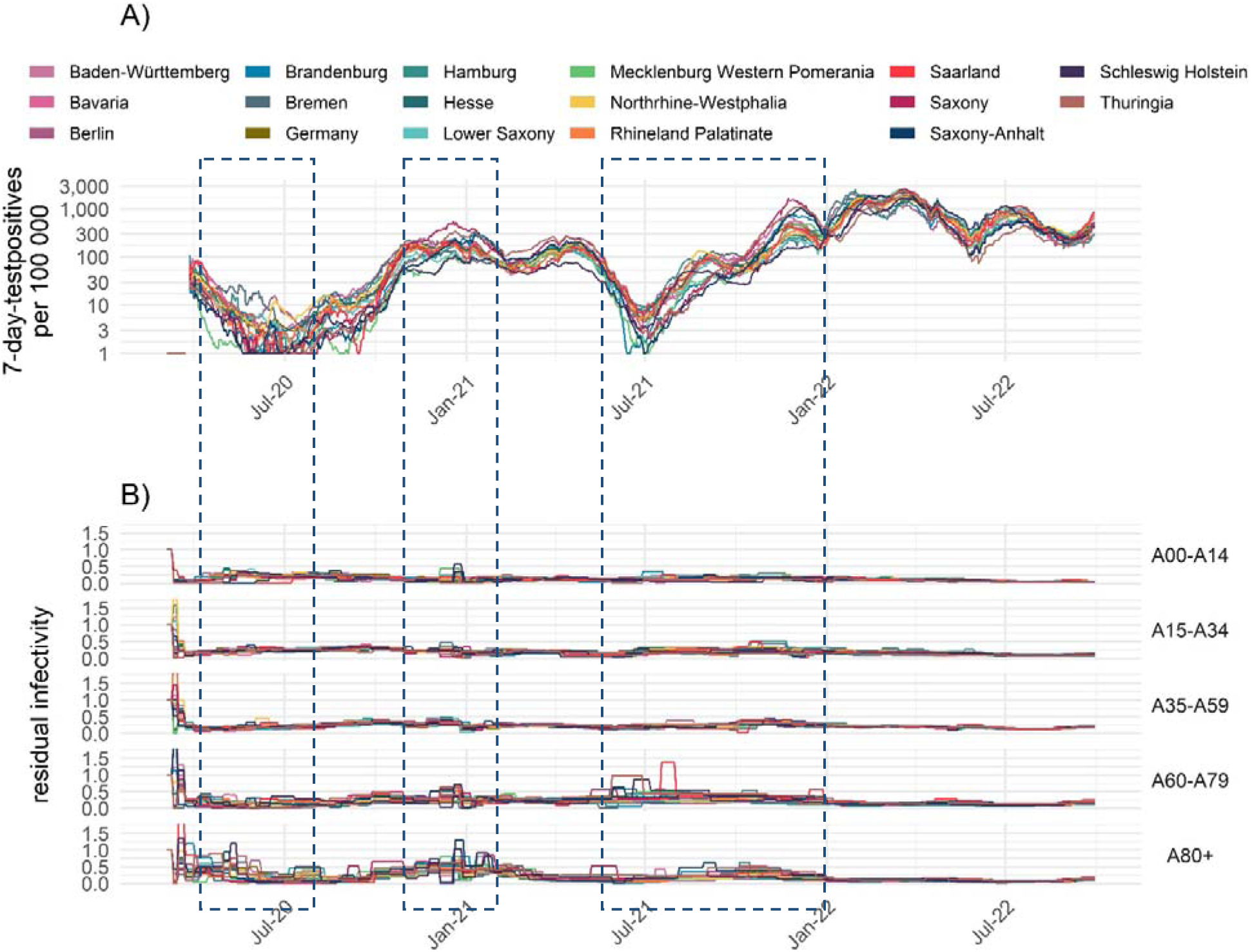
Comparison of between-state heterogeneity of infection dynamics, testpositivity rates and residual infectivity. A) Reported test-positives per region B) test positivity C) Estimated dynamical infecting rate b1. Time periods with increased heterogeneity between states are indicated with dashed rectangles. Here, heterogeneity is particularly high for both reported testpositivity and residual infectivity b1.

**Figure 7:**
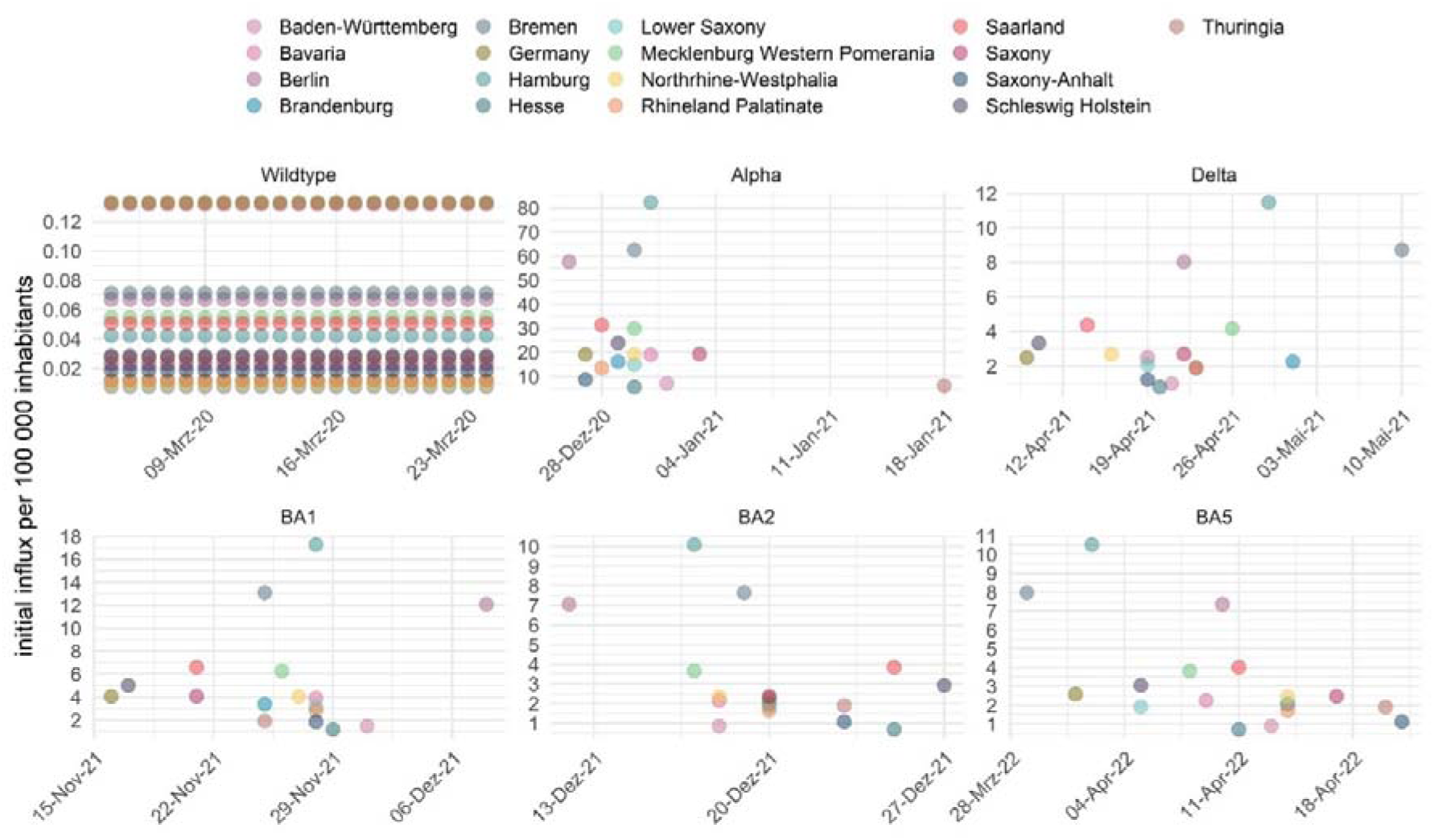
Region-specific model - estimation of the number and timepoints when successive SARS-Cov-II virus variants were introduced in each federal country

By estimating region-specific parameters as examplarily shown for the transfer rates from compartment infected I2 to compartment D dead the model can account for such differences (D). Consistently, federal countries Saxony and Thuringia, having suffered from high excess mortality in elderly in the alpha and delta wave, show highest parameter estimates in the oldest age group 80+. Regional specific parameter estimation for hospitalization rates and rates progressing to ICU are shown in Error! Reference source not found. and Error! Reference source not found., respectively

### Validated Model Predictions

We regularly used our model to predict scenarios of the future course of the epidemic and published these predictions via our website (XXX). We here present comparisons of our predictions with the actual course of the pandemic in order to validated our model.

### Comparsion of lockdowns December 2020 and November 2021 in Saxony

We used our model to predict the impact of lockdown measures on residual infectivities of our age groups. Comparing the lockdowns implemented in December 2020 and November 2021 in Saxony, it revealed that similar reductions were achieved except for the youngest age group (Figure 8A). Indeed, this is plausible because schools and day care facilities remained open in the November 2021 lockdown compared to the December 2020 lockdown.

**Figure 8:**
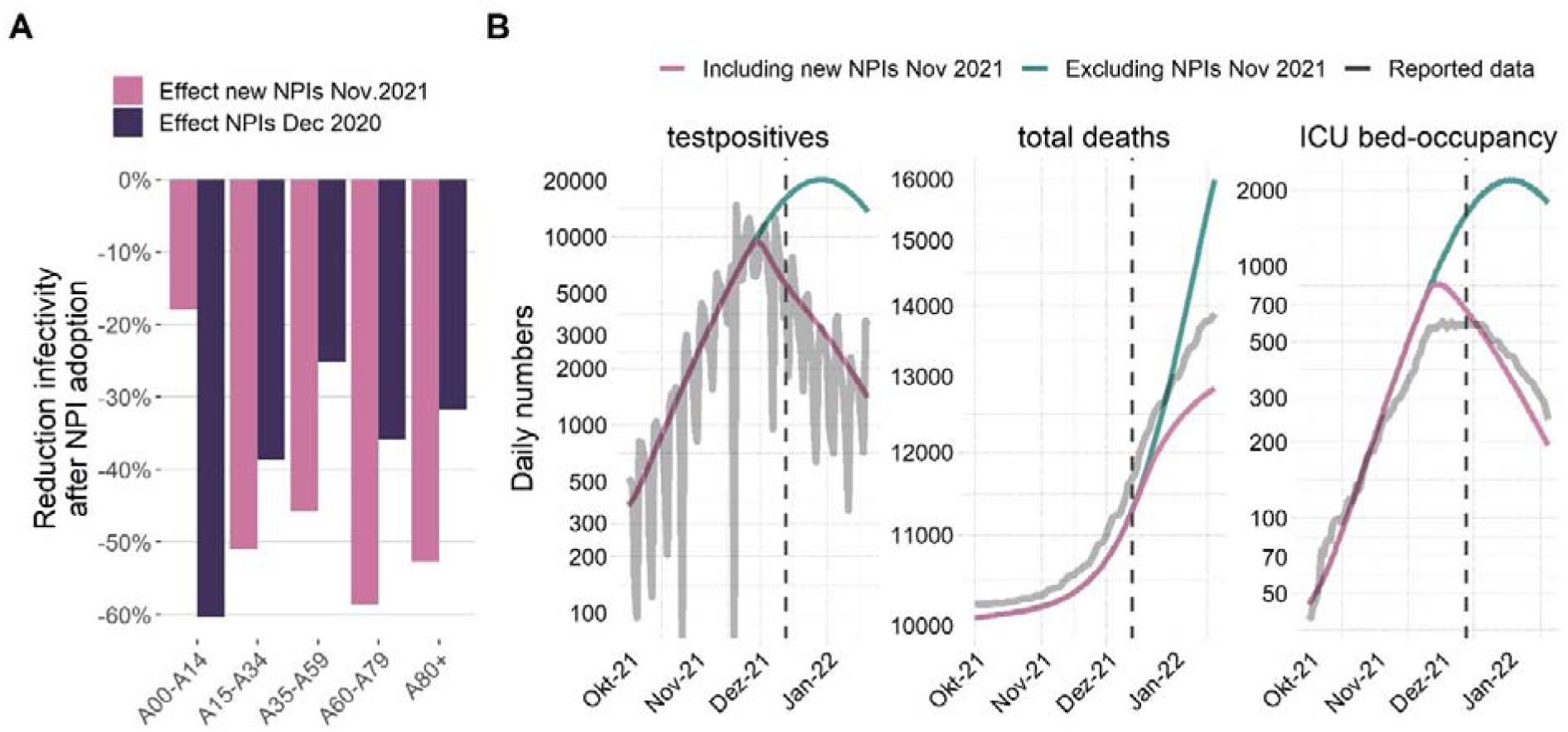
Estimation of the impact of the lockdown measures introduced in November 2021 in Saxony and compared to the December 2020 lockdown. A) Model-based estimate in decline of infectivity after the introduction of more stringent lockdown measures at December 2020 and November 2021 as a three-week average. While clear and comparable reductions in infectivities was estimated for the age-groups older than 14, the reduction for the younger age-group was much smaller in 2021 compared to 2020. This is plausible because schools and day care remained open in the 2021 lockdown. B) Modeled scenarios with and without introduction of lockdown. Without lockdown, the model predicted that infection numbers would increase until the end of December, and that the number of deaths would be increased in the order of >1,000 (https://www.health-atlas.de/documents/34). Model predictions are in reasonable agreement to the actual data. We used data from 2020-03-04 to 2021-12-13 to fit the model (dashed line). NPI: Non-pharmaceutical interventions

Based on these estimates, we predicted the further course of the pandemic after initiating the November 2021 lockdown and compared the results with a scenario without lockdown measures (Figure 8B). We estimated that the lockdown might have saved a four-digit number of lifes. In addition, the lockdown might have roughly halved the peak in ICU bed occupancy compared to the scenario without lockdown assuming that these ICU bed numbers could be supported. Finally, the further course of the pandemic closely resembles our model prediction for several weeks.

### Impact of higher vaccination rate

In Figure 9, we show the results of another modeled scenario to assess the impact of the higher vaccination rate in the federal state Saarland compared to the average German vaccination rate. We modeled the time course in Saarland in two scenarios, one with the actual vaccination rate and one with the reduced German vaccination rate. In the higher vaccination scenario, infections in the delta wave were significantly lower, while infections in the later omicron BA4+5 wave were significantly higher. This is likely due to the immune escape of the omicron variants regarding vaccination. However, the total number of deaths remained lower in the scenario observed with the higher vaccination rate, consistent with the reported lower pathogenicity of the Omicron variant. This suggests that the higher vaccination rate in Saarland may have played a relevant role in preventing severe outcomes also in the long term.

**Figure 9:**
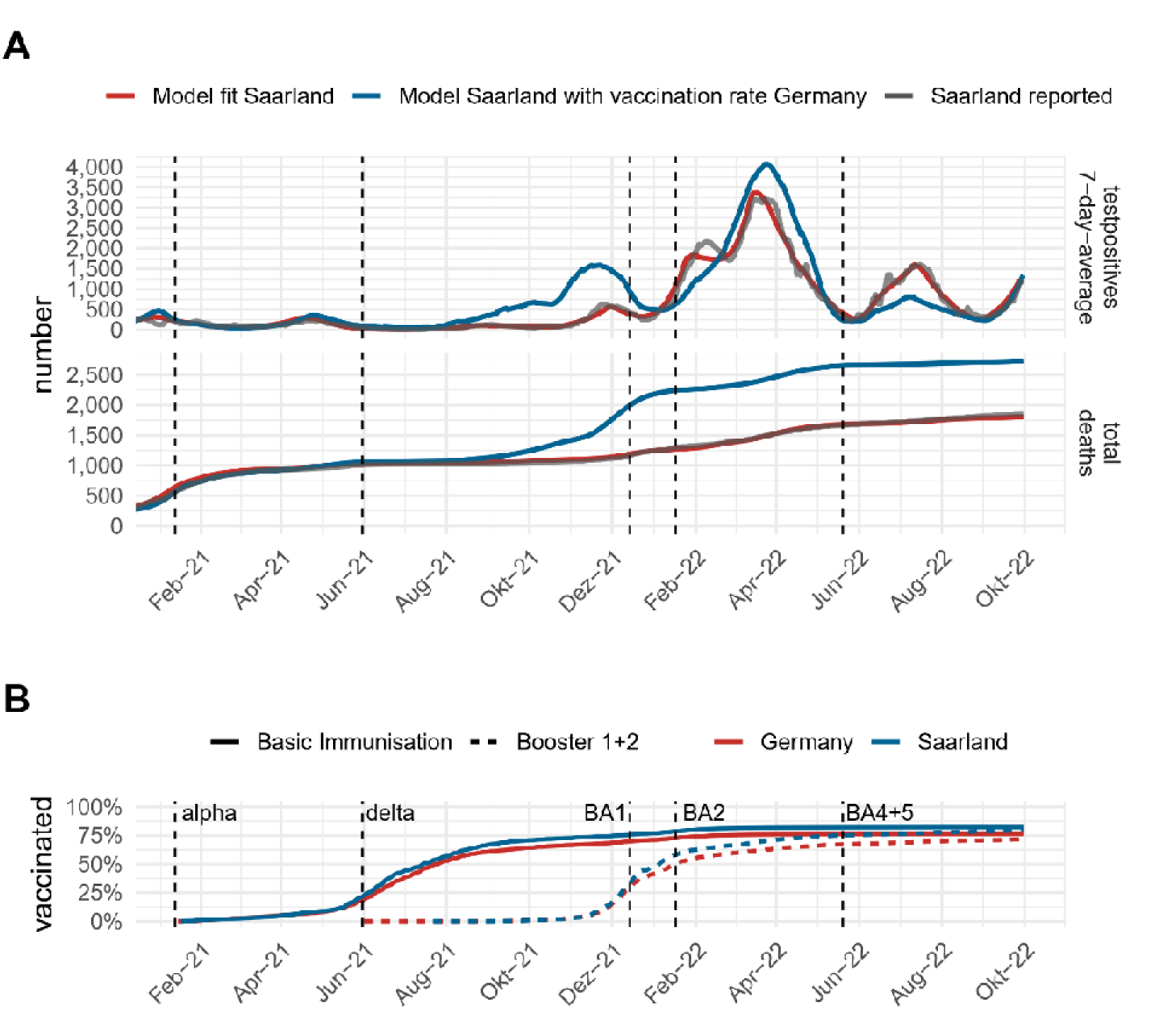
Estimation of the impact of the higher vaccination rate in the federal state Saarland compared with the German average. A) Comparison of the reported testpositives and COVID-19 deaths (grey) with the model under the observed vaccination rate (red). In blue, we show the hypothetical scenario of the lower average German vaccination rate applied in Saarland. Under this scenario, higher infection numbers of Omicron BA4+5 would be expected but still, death toll remains lower, B) Cumulative vaccination rates in Saarland compared with Germany. The occurrence of new SARS-CoV-II variants at frequencies higher than 5% is shown as dashed lines.

## Discussion

In this paper, we implement the early proposed method of parametrization of COVID-19 epidemiologic extended SECIR-type model [2] to explain the course of the epidemic in Germany as well as in all 16 German federal lands.

As in the previous case, we embed differential equations-based epidemic modelling into an input-output dynamical system (IO-NLDs), combining explicit mechanistic models of epidemic spread and phenomenological considerations of external impacts on model parameters via the input layer. As earlier, we assumed here also a non-direct link between state parameters of the embedded SECIR model and observables. This allows interposing a data model considering known biases of the available data resources.

Our current model updates the previous one by assuming age-dependent sub-models with known contact matrix and extending virus-variant dependent sub-models from two to unlimited (currently ten). We modelled vaccination, waning and boostering as well. The resulting updates enable us to get rid of two from three empirical dynamical parameters pcrit and pdeath, explaining variations in probability to develop critical symptoms or dye to age differences, different NPI for each age category and difference between virus variants. This shows, how unexplained empirical inputs can be later mechanically modelled, when additional information becomes available. This proofs universal importance of IO-NLDs, which enable to implement all current knowledge and ready update the model.

We model currently also hospital station occupancy. Along with ICU we model these outputs as an additional layer “hospitalization”, based indirectly on symptomatic compartments, rather than assuming separate model compartments.

Based on our IO-NLDS formulation and data models, we parametrized our model on the basis of data of infection numbers, critical and general hospital occupations, and deaths available for Germany and Saxony. Here, we chose a full-information approach considering all data in between start of the epidemic 4 March 2020 to 12 September 2024. We also applied a Bayesian learning process by considering other studies to inform model parameter’s settings. Thus, we combine mechanistic model assumptions with results from other studies and observational data. This approach is very popular in pharmacology [4] but despite its importance it is yet rarely applied in epidemiology [5].

Model parametrization resulted in a good and unbiased fit of data for the period considered for Germany and all its federal lands. Fixed parameter values of the SECIR model did not significantly deviated from known from literature values if available. A total of 37 intensification and relaxation events were necessary to describe the epidemic dynamics over the time course of observations for each federal subject and each age category, i.e. 85 different dynamical parameters of residual infectivity b1.

We estimated higher values of b1 at the very beginning of the epidemic, which could be due to natural contacting behaviour but could also be caused by issues regarding reporting or lack of testing capacities, i.e. we cannot exclude that this is an unresolved data artifact.

Seasonal changes can be discernable only in 2020. In 2021 the seasonality cannot be clearly traced. In the year 2021 the variability of b1 between federal states was large, exceeding variability among age groups for the overall data of Germany. This is plausible because NPI implementation was very heterogeneous between states. Only later in the pandemic (end of 2021), there were attempts to implement nation-wide harmonization of NPI rules and measures (see Error! Reference source not found.).

In 2022 b1 was mostly less than 0.2 in all age categories and stable with very little variability between federal states, showing effectivity of these nation-wide harmonization of NPI rules. Please note that the model’s predictions are based solely on data and assumptions. Therefore, the results should be interpreted with caution.

In this work we estimated 8917 parameters using more than 560000 data points: 1653 data points for 17 federal subjects for five age categories for new registered cases, death, hospital and ICU stations occupations as well as daily age-specific data on vaccination and waning.

Estimated infectivity roughly correlated with the Governmental Stringency Index [29]. We regularly contributed forecasts of our model to the German forecast Hub [27].

We also demonstrated utility of our model by several mid-term simulations of scenarios of epidemic development in Saxony, a federal state of Germany. We could show that predictions of reported infections were in the range of later observations for scenarios considered likely.

As future extensions and improvements of our model, we will consider stochastic effects on a daily scale, for example to model random influxes of cases or to model random extinctions of infection chains. These effects are relevant to be considered in times of low incidence numbers such as those observed in Germany in the summers 2020 and 2021. Our IO-NLDS framework is well suited to implement such extensions [3]. Furthermore, another application of our model is to provide infection scenarios for hospital data exposure models as previously successfully applied for german ICU admission forecasts or similar hospital resource planning tools currently being developed. This leveragaes the advantage of our modell, that compartments reflecting severe course are modeled solely as output- compartments, without serving as input for other model compartments.

In summary, the primary focus of the paper is an adequate parametrization of epidemiological models on the basis of complex, possibly biased data, as well as its coupling with structurally unknown dynamical external influences. This approach allows for a clear separation of mechanistic model compartments from random or time-dependent non-mechanistic influences and biases in the data. We believe that this approach is useful not only for the parametrization of the SECIR model presented here but also for other epidemiologic models including other disease contexts and data structures.

## Author Contributions

Conceptualization (ideas; formulation or evolution of overarching research goals and aims): H.K. and M.S.; data curation (management activities to annotate (produce metadata), scrub data and maintain research data (including software code, where it is necessary for interpreting the data itself) for initial use and later reuse): H.K.; formal analysis (application of statistical, mathematical, computational, or other formal techniques to analyze or synthesize study data): Y.K.; funding acquisition (acquisition of the financial support for the project leading to this publication): A.S. and M.S.; investigation (conducting a research and investigation process, specifically performing the experiments, or data/evidence collection): Y.K., H.K., and M.S.; methodology (development or design of methodology; creation of models): Y.K., M.S., and H.K.; project administration (management and coordination responsibility for the research activity planning and execution): A.S. and M.S.; resources (provision of study materials, reagents, materials, patients, laboratory samples, animals, instrumentation, computing resources, or other analysis tools): H.K.; software (programming, software development; designing computer programs; implementation of the computer code and supporting algorithms; testing of existing code components): Y.K. and H.K.; supervision (oversight and leadership responsibility for the research activity planning and execution, including mentorship external to the core team): M.S.; validation (verification, whether as a part of the activity or separate, of the overall replication/reproducibility of results/experiments and other research outputs): H.K. and M.S.; visualization (preparation, creation and/or presentation of the published work, specifically visualization/data presentation): H.K., writing—original draft preparation (creation and/or presentation of the published work, specifically writing the initial draft (including substantive translation)): Y.K., H.K., and M.S.; writing—review and editing (preparation, creation and/or presentation of the published work by those from the original research group, specifically critical review, commentary or revision—including pre- or post-publication stages): Y.K., H.K., A.S. and M.S. All authors have read and agreed to the published version of the manuscript.

## Institutional Review Board Statement

Ethical review and approval were waived for this study as only published data from official sources was used (see manuscript for detailed description of data sources).

## Informed Consent Statement

Patient consent was waived for this study as only published data from official sources was used (see manuscript for detailed description of data sources).

## Data Availability Statement

Code and data is available via GitHub at https://github.com/GenStatLeipzig/Modelling-dynamics-of-SARS-CoV-2-pandemics-of-Germany and in the Health Atlas https://www.health-atlas.de/models/40

## Acknowledgement

We thank Ministerial Director Daniel Stich from the Ministry of Science and Health Rhineland–Palatinate (MWG) for funding SentiSurv, Prof. Dr. Philipp Wild, Rieke Baumkötter, Simge Yilmaz, and the team from the University Medical Center Mainz for making the SentiSurv data available for our study, and the thousands of participants who contributed to public health through their commitment to the SentiSurv study

## Supplementary Titles

Appendix A Explanation of model structure, compartments and parameters Error! Bookmark not defined. General description of the model structure Error! Bookmark not defined.

Appendix B. Model equations Error! Bookmark not defined.

In this section, we present all equations of our model. Error! Bookmark not defined. Susceptible compartments Error! Bookmark not defined.

Sub-model of infected compartments Error! Bookmark not defined. Recovered compartments Error! Bookmark not defined. Appendix C. Input layer Error! Bookmark not defined. Appendix D. Output Layer Error! Bookmark not defined.

Appendix E. Parameter Estimation methods Error! Bookmark not defined.

Appendix F. Justification of fixed parameter values from the literature Error! Bookmark not defined. Appendix G. Estimation of unreported Cases (Dark Figure) Error! Bookmark not defined.

Appendix H. Agreement of model and data of reported cases for all federal states Error! Bookmark not defined. Appendix I. Agreement of model and data of severe disease courses per federal state Error! Bookmark not defined. Appendix J. Age-group specific dynamics of infectivity for Germany Error! Bookmark not defined.

Appendix K. Federal state specific parameter estimates of the transition rate from the infected compartment (I2) to the compartment of hospitalized cases (H) Error! Bookmark not defined.

Appendix L. Federal state specific parameter estimates of the transition rate from the infected compartment (I2) to the compartment of ICU cases (C) Error! Bookmark not defined.

Appendix M. Prediction of the dynamics of immune states for the federal states Error! Bookmark not defined.

Appendix N. Estimation of age-group specific temporal dynamics of immune states of infected subjects Error! Bookmark not defined.

## References

1. Kong L, Duan M, Shi J, Hong J, Chang Z, Zhang Z. Compartmental structures used in modeling COVID-19: a scoping review. Infect Dis Poverty. 2022; 11:72. Epub 2022/06/21. doi: 10.1186/s40249-022-01001-y PMID: 35729655.

2. Kheifetz Y, Kirsten H, Scholz M. On the Parametrization of Epidemiologic Models—Lessons from Modelling COVID-19 Epidemic. Viruses. 2022; 14. doi: 10.3390/v14071468 PMID: 35891447.

3. Georgatzis K, Williams CKI, Hawthorne C. Input-Output Non-Linear Dynamical Systems applied to Physiological Condition Monitoring. Proceedings of the 1st Machine Learning for Healthcare Conference 2016: PMLR; 19 August 2016 through 20 August 2016 [updated 2016 Aug 19 through 2016 Aug 20].

4. Friberg LE, Henningsson A, Maas H, Nguyen L, Karlsson MO. Model of chemotherapy-induced myelosuppression with parameter consistency across drugs. J Clin Oncol. 2002; 20:4713–21. doi: 10.1200/JCO.2002.02.140 PMID: 12488418.

5. Dehning J, Zierenberg J, Spitzner FP, Wibral M, Neto JP, Wilczek M, et al. Inferring change points in the spread of COVID-19 reveals the effectiveness of interventions. Science. 2020; 369. Epub 2020/05/15. doi: 10.1126/science.abb9789 PMID: 32414780.

6. Schuppert A, Theisen S, Fränkel P, Weber-Carstens S, Karagiannidis C. Bundesweites Belastungsmodell für Intensivstationen durch COVID-19. Med Klin Intensivmed Notfmed. 2021. Epub 2021/02/03. doi: 10.1007/s00063-021-00791-7 PMID: 33533980.

7. Mossong J, Hens N, Jit M, Beutels P, Auranen K, Mikolajczyk R, et al. Social contacts and mixing patterns relevant to the spread of infectious diseases. PLoS Med. 2008; 5:e74. doi: 10.1371/journal.pmed.0050074 PMID: 18366252.

8. Der Heiden M an, Buchholz U. Modellierung von Beispielszenarien der SARS-CoV-2-Ausbreitung und Schwere in Deutschland. Robert Koch-Institut; 2020.

9. Nishiura H, Linton NM, Akhmetzhanov AR. Serial interval of novel coronavirus (COVID-19) infections. Int J Infect Dis. 2020; 93:284–6. Epub 2020/03/04. doi: 10.1016/j.ijid.2020.02.060 PMID: 32145466.

10. Tindale LC, Stockdale JE, Coombe M, Garlock ES, Lau WYV, Saraswat M, et al. Evidence for transmission of COVID-19 prior to symptom onset. Elife. 2020; 9. Epub 2020/06/22. doi: 10.7554/eLife.57149 PMID: 32568070.

11. Böhmer MM, Buchholz U, Corman VM, Hoch M, Katz K, Marosevic DV, et al. Investigation of a COVID-19 outbreak in Germany resulting from a single travel-associated primary case: a case series. The Lancet Infectious Diseases. 2020; 20:920–8. doi: 10.1016/S1473-3099(20)30314-5.

12. Ganyani T, Kremer C, Chen D, Torneri A, Faes C, Wallinga J, et al. Estimating the generation interval for coronavirus disease (COVID-19) based on symptom onset data, March 2020. Euro Surveill. 2020; 25. doi: 10.2807/1560-7917.ES.2020.25.17.2000257 PMID: 32372755.

13. Zhou F, Yu T, Du R, Fan G, Liu Y, Liu Z, et al. Clinical course and risk factors for mortality of adult inpatients with COVID-19 in Wuhan, China: a retrospective cohort study. The Lancet. 2020; 395:1054–62. doi: 10.1016/S0140-6736(20)30566-3.

14. Sanche S, Lin YT, Xu C, Romero-Severson E, Hengartner N, Ke R. High Contagiousness and Rapid Spread of Severe Acute Respiratory Syndrome Coronavirus 2. Emerg Infect Dis. 2020; 26:1470– 7. Epub 2020/06/21. doi: 10.3201/eid2607.200282 PMID: 32255761.

15. COVID-19 National Emergency Response Center. Coronavirus Disease-19: The First 7,755 Cases in the Republic of Korea. Osong Public Health Res Perspect. 2020; 11:85–90. doi: 10.24171/j.phrp.2020.11.2.05 PMID: 32257774.

16. Tolksdorf K, Buda S, Schuler E, Wieler LH, Haas W. Eine höhere Letalität und lange Beatmungsdauer unterscheiden COVID-19 von schwer verlaufenden Atemwegsinfektionen in Grippewellen. 2020. Epub 2020/08/28. doi: 10.25646/7111.

17. Karagiannidis C, Mostert C, Hentschker C, Voshaar T, Malzahn J, Schillinger G, et al. Case characteristics, resource use, and outcomes of 10 021 patients with COVID-19 admitted to 920 German hospitals: an observational study. The Lancet Respiratory Medicine. 2020; 8:853–62. doi: 10.1016/S2213-2600(20)30316-7.

18. Linton NM, Kobayashi T, Yang Y, Hayashi K, Akhmetzhanov AR, Jung S-M, et al. Incubation Period and Other Epidemiological Characteristics of 2019 Novel Coronavirus Infections with Right Truncation: A Statistical Analysis of Publicly Available Case Data. J Clin Med. 2020; 9. Epub 2020/02/17. doi: 10.3390/jcm9020538. PMID: 32079150.

19. Verity R, Okell LC, Dorigatti I, Winskill P, Whittaker C, Imai N, et al. Estimates of the severity of coronavirus disease 2019: a model-based analysis. The Lancet Infectious Diseases. 2020; 20:669–77. doi: 10.1016/S1473-3099(20)30243-7.

20. Bock W, Jayathunga Y, Götz T, Rockenfeller R. Are the upper bounds for new SARS-CoV-2 infections in Germany useful. Computational and Mathematical Biophysics. 2021; 9:242–60. doi: 10.1515/cmb-2020-0126.

21. https://www.divi.de/divi-intensivregister-tagesreport-Tagesreport-Archiv. Tagesreport-Archiv. DIVI 2020 [updated 30 Jun 2022].

22. Robert-Koch Institut, editor. COVID-19 Hospitalisierungen in Deutschland. 2024. Available from: https://github.com/robert-koch-institut/COVID-19-Hospitalisierungen_in_Deutschland.

23. Robert Koch-Institut. SARS-CoV-2 Sequenzdaten aus Deutschland. Zenodo; 2023.

24. Bericht zu Virusvarianten von SARS-CoV-2 in Deutschland, insbesondere zur Variant of Concern (VOC) B.1.1.7. Robert Koch Institute [updated 3 Mar 2021; cited 3 Mar 2021]. Available from: https://www.rki.de/DE/Content/InfAZ/N/Neuartiges_Coronavirus/DESH/Bericht_VOC_2021-03-03.pdf?__blob=publicationFile.

25. Kheifetz Y, Scholz M. Modeling individual time courses of thrombopoiesis during multi-cyclic chemotherapy. PLoS Comput Biol. 2019; 15:e1006775. Epub 2019/03/06. doi: 10.1371/journal.pcbi.1006775 PMID: 30840616.

26. Advances in neural information processing systems. ; 1990.

27. Hooke R, Jeeves TA. “ Direct Search” Solution of Numerical and Statistical Problems. J ACM. 1961; 8:212–29. doi: 10.1145/321062.321069.

28. Kreutz C, Raue A, Kaschek D, Timmer J. Profile likelihood in systems biology. FEBS J. 2013; 280:2564–71. Epub 2013/05/09. doi: 10.1111/febs.12276 PMID: 23581573.

29. https://www.bsg.ox.ac.uk/research/research-projects/covid-19-government-response-tracker. COVID-19 Government Response Tracker. University of Oxford 2020 [updated 3 Mar 2022].

